# Safety and Immunogenicity of Biological E’s 14-valent Pneumococcal Conjugate Vaccine (PNEUBEVAX 14^®^) Administered in a 3p+1 Schedule to Healthy Indian Infants and Toddlers: A Prospective, Multicenter, Active Controlled Phase IV Trial

**DOI:** 10.64898/2025.12.18.25342547

**Authors:** Subhash Thuluva, Ramesh V. Matur, Subbareddy Gunneri, Siddalingaiah Ningaiah, Vijay Yerroju, Rammohan Reddy Mogulla, Chirag Dhar, Kamal Thammireddy, Shivani Desai, Piyush Paliwal, Raju Esanakarra, Spoorthy Narayandas, B. S. Chakravarthy, Jai Prakash Narayan, N. S. Mahantshetti, Manish Narang, Ramanath Andy Karayar, Savita Verma, Pareshkumar. A. Thakkar, Jog Pramod Prabhakar, Safety & Antibody Assessments THrough Routine Immunization with PNEUBEVAX 14® (SAATHI-14) investigators

**Affiliations:** R&D - Clinical Development, Biological E. Limited, Hyderabad, India; Clinical Serology, Biological E. Limited, Hyderabad, India; King George Hospital, Vishakhapatnam, India; JLN Medical College, Ajmer, India; KLES Dr. Prabhakar Kore Hospital & Medical Research Centre, Belagavi, India; GTB Hospital, Delhi, India; Panimalar Medical College Hospital & Research Institute, Chennai, India; Pt. B D Sharma Post Graduate Institute of Medical Sciences & Hospital, Rohtak, India; SSG General Hospital. Vadodara, India; Medipoint Hospital, Pune, India

## Abstract

**Background:** Biological E’s PNEUBEVAX 14^®^ (BE-PCV14) is a WHO-prequalified 14-valent pneumococcal conjugate vaccine that adds emerging serotypes 22F and 33F to PCV13 coverage. We compared the safety and immunogenicity of a 3p+1 infant schedule of BE-PCV14 versus Prevenar 13^®^ (PCV13) in Indian infants.

**Methods:** In this prospective, open-label, multicentre phase IV trial, PCV-naïve infants aged 6–8 weeks received three primary doses of BE-PCV14 or PCV13 followed by a booster at 12–15 months. In the immunogenicity subset, serotype-specific IgG to the 14 vaccine serotypes and cross-protective 6A was measured 28 days post-booster. Endpoints were seroresponse rates (SRR defined as IgG ≥ 0.35 µg/mL), IgG geometric mean concentrations (GMCs), and frequency of solicited / unsolicited adverse events (AEs) / serious AEs (SAEs) through 28 days post-booster. *Post hoc* non-inferiority of BE-PCV14 to PCV13 for all 14 serotypes was assessed by calculating the SRR differences and GMC ratios. For the SRR difference, non-inferiority was to be concluded if the lower limit of the 95% confidence interval was >-10%, and for the GMC ratio if the lower limit of the 95% confidence interval was >0.5.

**Results:** Post-booster immunogenicity was assessed in 559 BE-PCV14 and 147 PCV13 recipients. AEs occurred at similarly low frequencies in both groups (∼6%), were mainly mild local reactions or pyrexia, and the two SAEs after BE-PCV14 were considered unrelated to vaccination. Post-booster SRRs with BE-PCV14 were ≥92.8% for all 14 serotypes and >90% for 6A, comparable to PCV13 for shared serotypes. BE-PCV14 induced strong responses to the additional serotypes 22F and 33F, and post-booster/pre-booster GMC ratios were approximately 2.4–5.3 across serotypes. *Post hoc* analyses showed BE-PCV14 was non-inferior to PCV13 for all 14 serotypes by both SRR and GMC criteria.

**Conclusions:** When used as a booster in a 3p+1 schedule, PNEUBEVAX 14^®^ is well tolerated and elicits robust booster responses comparable to PCV13 while extending serotype coverage to 15 serotypes, including emerging 22F and 33F and cross-protective 6A.

## Introduction

Worldwide, *Streptococcus pneumoniae* is a leading cause of pneumonia in paediatric and adult populations, with majority of cases in low- and middle-income countries (LMICs). The World Health Organization (WHO) reports that 14% of global deaths in children under 5 years of age are due to pneumonia, which is higher than deaths due to malaria, AIDS, and measles combined **(1)**. While pneumonia affects children globally, most deaths occur in South Asia and sub-Saharan Africa **(2,3)**. In addition to pneumonia, *Streptococcus pneumoniae* also contributes to invasive infections such as meningitis and sepsis, which are less common but more severe. Acute otitis media, which occurs more commonly than invasive disease, is also often caused by *Streptococcus pneumoniae.* To mitigate the burden of these diseases, the WHO promotes pneumococcal conjugate vaccines (PCVs) as an important public health tool to be incorporated into national immunization programs globally **(4)**. While the WHO recommends a 3-dose series, the Indian Academy of Paediatrics’ (IAP) Advisory Committee on Vaccines & Immunization Practices (ACVIP) recommends a 4-dose series (3p+1) **(5)**. The introduction of 7-, 10- and 13-valent PCVs into infant immunization programmes has considerably reduced the burden of invasive pneumococcal disease (IPD) and pneumonia **(6,7)**. However, non-vaccine serotypes have emerged in circulation **(8)**, with serotypes 22F and 33F being the prevalent ones **(9,10)**. The emergence underscores the need for vaccines that extend serotype coverage, such as Biological E’s PNEUBEVAX 14^®^ (BE-PCV14), a 14-valent PCV that includes serotypes 22F and 33F. BE-PCV14 underwent a robust clinical development program that led to its recent prequalification by the WHO. Clinical data from a Phase I, Phase II, and three Phase III clinical trials demonstrated the safety and robust immunogenicity of BE-PCV14. The pivotal Phase III clinical trial demonstrated BE-PCV14’s immunogenic non-inferiority to PCV13 with a comparable safety profile **(11)**. Two other Phase III trials demonstrated the safety and immunogenicity of the multi-human dose formulation of BE-PCV14 **(12)**, as well as its safety and immunogenicity in a 2p+1 schedule **(13)**. Both trials also demonstrated *post hoc* immunogenic non-inferiority to PCV13. While BE-PCV14 has shown strong evidence of safety and immunogenicity in a 3p+0 and 2p+1 schedule, an important question remained: BE-PCV14’s profile in a 3p+1 schedule. Countries such as Japan and the USA use this infant/toddler immunization schedule **(14)**. Guided by updated evidence on PCVs, the IAP’s ACVIP also recommends this schedule **(15)**. As part of a large Phase IV post-marketing study of BE-PCV14, the 3p+1 schedule was studied in comparison with PCV13. Here, we report the results on the safety and immunogenicity of a BE-PCV14 booster in a 3p+1 schedule.

## Materials and Methods

### Study Design and participants

SAATHI-14 (<u>S</u>afety and <u>A</u>ntibody <u>A</u>ssessment <u>Th</u>rough Routine Immunization with PNEUBEVAX <u>14</u>^®^) was a prospective, multicentre, phase IV study conducted at 31 sites within India to evaluate the safety and immunogenicity of Biological E’s 14-valent pneumococcal conjugate vaccine PNEUBEVAX 14^®^ (BE-PCV14). A total of 2600 healthy eligible infants (6–8 weeks old at the time of first dose administration) of either sex were recruited at the study sites. Among the total enrolled participants, 2300 were enrolled under BE-PCV14 and 300 under PCV13. Key eligibility criteria are summarized in Appendix 1 of the supplementary information. Within this study, a subset was selected to evaluate the safety and immunogenicity of a three-dose primary series followed by a booster dose (3p+1) schedule of BE-PCV14. The study assessed serotype-specific IgG immune responses against all 14 serotypes of BE-PCV14. All SAATHI-14 study sites and principal investigators are listed in Appendix 2 of the supplementary information.

The study was conducted in two parallel cohorts to comprehensively evaluate the safety and immunogenicity of a booster of BE-PCV14 in toddlers. Figure 1 represents the overall study design of the trial. The first cohort focused on safety assessment and enrolled 1400 participants, who received three 0.5 mL intramuscular doses of BE-PCV14 administered 28 days apart. The study vaccine was co-administered with routine childhood immunizations, including DTwP-HepB-Hib-IPV and rotavirus vaccines, in accordance with the national immunization schedule. A booster dose was administered for all available consented study participants at 12–15 months of age as per their study vaccine assignment to assess safety and immunogenicity. The second cohort comprised 1200 participants and was designed to evaluate both immunogenicity and safety. Participants in this cohort were allocated to receive either BE-PCV14 (n=900) or a licensed comparator, PCV13 (n=300). Of the n=900 toddlers eligible to receive a booster, n=600 were to be enrolled to receive a BE-PCV14 booster dose and return for a post-booster immunogenicity visit. Of these, n=100 were also to have a pre-booster immunogenicity blood draw. In parallel, all n=300 who received the primary series of PCV13 were to be enrolled to receive a booster and a subsequent post-booster immunogenicity visit with n=50 of these having a pre-booster blood draw.

**Figure 1:**
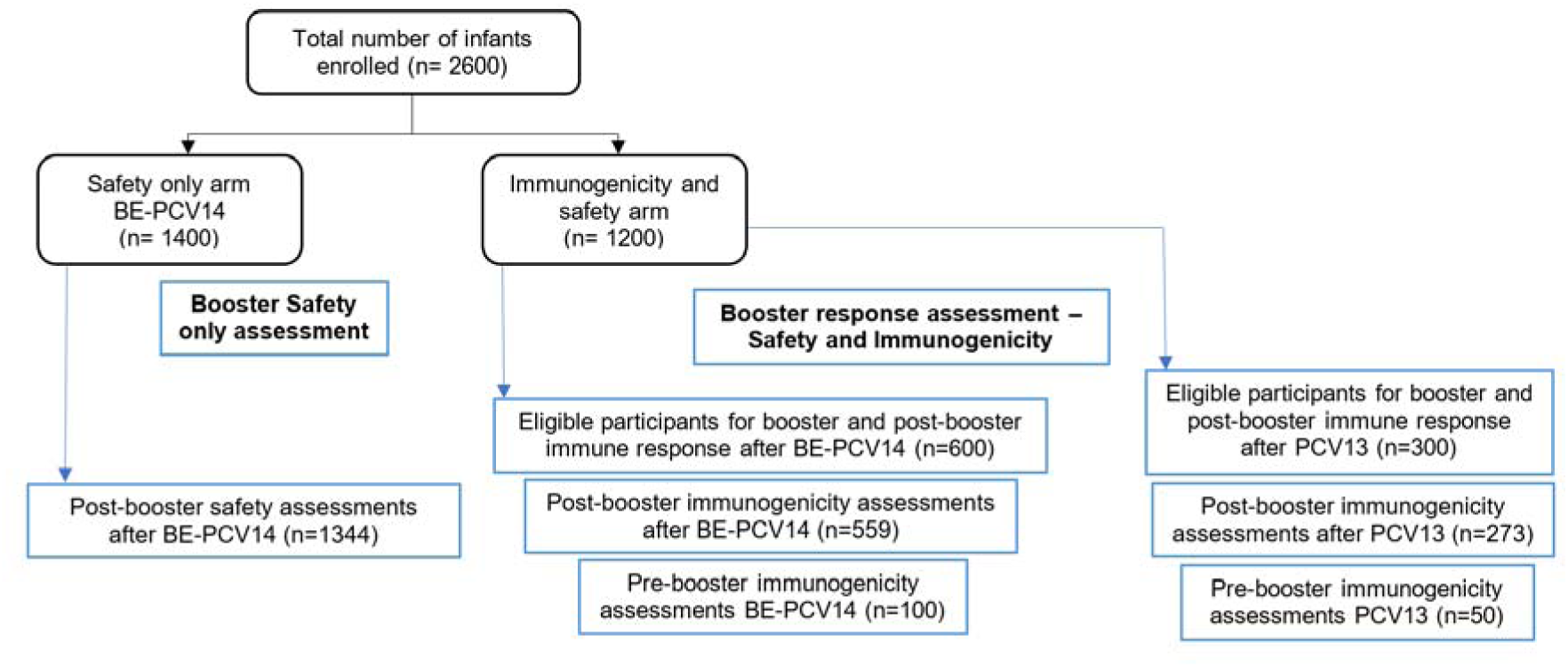
Study Schema.

### Ethical considerations

The study protocol was approved by the institutional ethics committees of all 31 SAATHI-14 sites listed in the supplementary information. Additionally, the protocol was approved by the Central Drugs Standard Control Organisation (CDSCO), the Indian National Regulatory Agency and registered prospectively on ctri.nic.in (CTRI/2023/09/057894). Informed written consent was sought and received from the parents of all participants in this study. This study was conducted in accordance with the Declaration of Helsinki and applicable ICH-GCP guidelines in effect at the time of the study.

### Randomization and masking

This booster study included both randomized and non-randomized participants in the final analyses. Briefly, N = 600 were randomized 1:1 on first enrolment to receive a 3-dose primary series of BE-PCV14 or PCV13. Apart from these participants, additional immunogenicity data were drawn from those that were enrolled for lot-to-lot consistency studies of BE-PCV14 per regulatory guidance. Additional booster safety data was collected from participants who were enrolled for the BE-PCV14 3-dose primary series safety study. A schematic of the study design is provided in Figure 1. This study was open-label, meaning both the parent(s)/LAR(s) of the participants, and the investigators were aware of which study vaccine the participant received. Laboratory personnel performing the immunological assays were blinded to sample identity.

### Procedures

The screening assessments included demographic data, medical history, general examination, and a complete systemic physical examination. The overall study consisted of multiple visits to receive the primary series of PCV. Relevant visits to this manuscript were the Day 84 post-primary immunogenicity visit, a booster visit at 12-15 months of age, and a post-booster safety and immunogenicity visit 28 days after. Here we report the data for the 3p+1 schedule, i.e., the safety and immunogenicity data until 28 days after the administration of a booster dose.

A total of 5.0 mL whole blood was collected by a trained phlebotomist three times during the study, once 28 days after the three primary doses (post-primary timepoint), again on the day the participant returned for a booster dose (pre-booster timepoint), and finally 28 days after the booster dose (post-booster timepoint). These samples were used for the analysis of the parameters as defined. Serum was separated by centrifugation at 2500 rpm for 5 –15 minutes at the study central laboratory. The separated serum was aspirated and transferred into two pre-labelled supplied cryovials and placed at -20 to -80°C.

Anti-pneumococcal capsular polysaccharide (anti-PnCPS) immunoglobulin G (IgG) antibody concentration estimation against each of the 14 vaccine serotypes and cross-reactive serotype 6A was performed as per the World Health Organization (WHO) reference ELISA with a minor modification. Instead of cell wall polysaccharide (CWPS), CWPS-multi was used in the assay. Also, 22F pneumococcal polysaccharide (PnPS) was used for pre-adsorption to neutralize non-specific antibodies and other common contaminants present in the PnPS coating antigens.

### Vaccines and Immunization procedures

BE-PCV14 (PNEUBEVAX 14^®^, Biological E. Limited, India) is a 14-valent pneumococcal polysaccharide conjugate vaccine containing capsular polysaccharides from *S. pneumoniae* serotypes 1, 3, 4, 5, 6B, 7F, 9V, 14, 18C, 19A, 19F, 22F, 23F and 33F, each conjugated to CRM_197_. For the multi-human dose presentation used in this study, each 0.5 mL dose contains 3.0 µg of polysaccharide for serotype 1, 2.2 µg for serotypes 3, 4, 5, 7F, 9V, 14, 18C, 19A, 19F, 22F, 23F and 33F, and 4.4 µg for serotype 6B, adsorbed onto ≤0.75 mg aluminium (as aluminium phosphate), with 4 mg 2-phenoxyethanol as preservative and the excipients polysorbate 20 and succinic acid.

PCV13 (PREVENAR 13^®^, Pfizer Inc., USA) consists of a 13-valent pneumococcal polysaccharide conjugate vaccine containing capsular polysaccharides from Streptococcus pneumoniae serotypes 1, 3, 4, 5, 6A, 6B, 7F, 9V, 14, 18C, 19A, 19F and 23F, each individually conjugated to the diphtheria CRM_197_ carrier protein. Each 0.5 mL paediatric dose in a pre-filled syringe contains 2.2 µg of polysaccharide for serotypes 1, 3, 4, 5, 6A, 7F, 9V, 14, 18C, 19A, 19F and 23F and 4.4 µg for serotype 6B, together with ∼32 µg CRM, 0.125 mg aluminium (as aluminium phosphate), and the excipients polysorbate 80, succinic acid, sodium chloride and water for injection.

The primary schedules for BE-PCV14 or PCV13 were administered as 0.5 mL intramuscular injection in one limb (anterolateral aspect of the thigh) and the other EPI vaccines (a 0.5 mL hexavalent vaccine [DTwP-rHepB-Hib-IPV]), co-administered intramuscularly on the other limb. A 0.5 mL rotavirus vaccine was co-administered orally in the same 6–10–14-week dosing schedule in both safety and immunogenicity arms. BE-PCV14 and PCV13 vaccines were then administered as a booster dose at 12–15 months of participant age. All the participants were observed for at least 30 minutes after administration of the booster to evaluate and treat any immediate adverse reactions. A diary card was provided to the participant’s parent/LAR to record any solicited local/general AEs occurring after vaccination. Solicited local and systemic AEs were collected during the 7 days (Day 0 to Day 6) after booster administration and unsolicited AEs, if any, until the end of the study 28 days after the booster. All serious AEs related to study participation were reported until the end of the study. All AEs and serious AEs (SAEs), whether leading to withdrawal from the study or not, were also reported/recorded until the end of the study.

### Outcomes

The outcome of the study was the assessment of the safety and immunogenicity of BE-PCV14 when given in a 3p+1 schedule. The safety endpoints included evaluation of solicited local and systemic AEs in participants during the first 30 minutes of the post-booster observation period and for the subsequent seven consecutive days (Day 0–6) thereafter, captured through a participant diary, unsolicited AEs during the 28 days post-booster dose follow-up period, and SAEs during the 28 days post-booster dose follow-up period.

The immunogenicity endpoints included descriptive comparison of anti-PnCPS IgG GMCs in BE-PCV14 vaccinated participants at 28 days post booster dose with GMCs at 28 days post 3-dose primary series for all 14 serotypes, anti-PnCPS IgG GMC ratio (post-boost GMC/post-primary GMC) for all 14 serotypes in participants vaccinated with BE-PCV14, and assessment of anti-PnCPS IgG GMCs and their ratios (post-booster GMC/pre-booster GMC) for all 14 serotypes from pre-booster to 28 days post booster dose immune response. A descriptive comparison of anti-PnCPS IgG GMCs against serotype 6A in BE-PCV14 and PCV-13 vaccinated participants at 28 days post booster dose with GMCs at 28 days post 3-dose primary series was another immunogenicity assessment. Post-booster seroresponse rates (defined as percentage of participants achieving a post-booster serotype-specific anti-PnCPS IgG concentration ≥0.35 µg/mL) and anti-PnCPS IgG GMC in the BE-PCV14 and PCV13 groups were also descriptively compared.

### Sample Size determination

In a pivotal Phase III trial of BE-PCV14 we had already demonstrated non-inferiority to PCV13 leading to the marketing authorization and subsequent WHO prequalification of BE-PCV14. The booster-response objectives of this Phase IV study were mainly to generate additional post-marketing evidence to support clinical decision-making for use of BE-PCV14 in a 3p+1 schedule. To generate this evidence, a sample size of N = 600 (N= 540 with a 10% dropout rate) for BE-PCV14 and N = 300 (N = 270 with a 10% dropout rate) was deemed to be sufficient to descriptively compare the safety and immunogenicity of the two vaccines. This number was based on pre-trial regulatory guidance from CDSCO, the Indian National Regulatory Agency.

### Statistical Analyses

Summary statistics were used wherever necessary. Visual data representations such as trellis plots and forest plots are included wherever appropriate. Occurrence rates of AEs / SAEs were compared descriptively and presented tabularly. Serotype-specific anti-pneumococcal capsular polysaccharide (PnCPS) IgG concentrations (µg/mL) were quantified in each vaccine group using a validated enzyme-linked immunosorbent assay (ELISA). The ratio of anti-PnCPS IgG GMC at 28 days post booster dose to 28 days post primary series along with their two-sided 95% CI was calculated. The ratio of anti-PnCPS IgG GMC 28-days post-booster dose to pre-booster GMC along with their 2-sided 95% CI was also calculated.

*Post hoc* non-inferiority analyses were performed to compare the post-booster responses of BE-PCV14 with PCV13. The difference in seroresponse rate and 95% CI was calculated for each vaccine serotype by the Farrington-Manning method. Non-inferiority was to be concluded if the lower limit of the 95% CI was >-10%. Similarly, post-booster GMC ratio and 95% CI (log-transformed and back transformed) of BE-PCV14 to PCV13 was also calculated for each vaccine serotype. Non-inferiority was to be concluded if the lower limit of the 95% CI of the ratio was >0.5.

## Results

### Study population and study disposition

In this Phase IV study, a total of 2605 healthy infants aged between 6–8 weeks of age were screened and 2600 were enrolled. There were four screen failures, and the parents of one participant withdrew consent after randomization. Of those who received the primary series of BE-PCV14, 1903 (82.7%) returned and received a booster dose. Of those who received PCV13, 273 returned for a booster dose. These subsets form the safety population. From within this safety population, 100 and 559 infants who received BE-PCV14, and 50 infants and 147 infants who received PCV13 were included for a pre-booster and post-booster blood draw respectively. This information and baseline clinical characteristics are provided in Table 1 and the study schema is presented in Figure 1.

**Table 1:** Baseline clinical characteristics.

| Characteristic | BE-PCV14 | PCV13 |
| --- | --- | --- |
| Total number of infants in study | N = 2300 | N = 300 |
| Mean Age (± SD) in days | 47.6 (± 3.95) | 48.2 (± 3.80) |
| Mean Length (± SD) in centimeters | 54.11 (± 2.09) | 53.68 (± 1.46) |
| Median Weight (Range) in kilograms | 4.046 (3.300 – 6.040) | 4.100 (3.300 – 6.100) |
| Total number of female infants (%) | n = 1013 (44%) | n = 126 (42%) |
| Number of participants that returned for a booster dose (safety population) | n = 1903 (82.7%) | n = 273 (91%) |
| Number of infants with a pre-booster immunogenicity visit | n = 100 | n = 50 |
| Number of infants with post-primary and post-booster immunogenicity visits | n = 559 | n = 147 |

### Safety findings

Overall, 147 adverse events were reported in the 119 children (6.25%) who received a BE-PCV14 booster dose. After a PCV13 booster dose, 16 AEs were reported in 16 children (∼6%). AEs in both study arms were largely solicited with the most frequent ones being injection site erythema, injection site pain, injection site swelling, and pyrexia. There were 9 unsolicited AEs reported in 5 children who received a BE-PCV14 booster dose and there were 2 children with serious AE (SAE) reports. One SAE of febrile seizure occurred in a study participant five days after receiving a booster dose, and another was a case of acute gastroenteritis about 14 days after receiving a booster dose. Both SAEs were adjudicated to be unrelated to the study vaccine by the respective principal investigators. Data are summarized in Table 2.

**Table 2:**
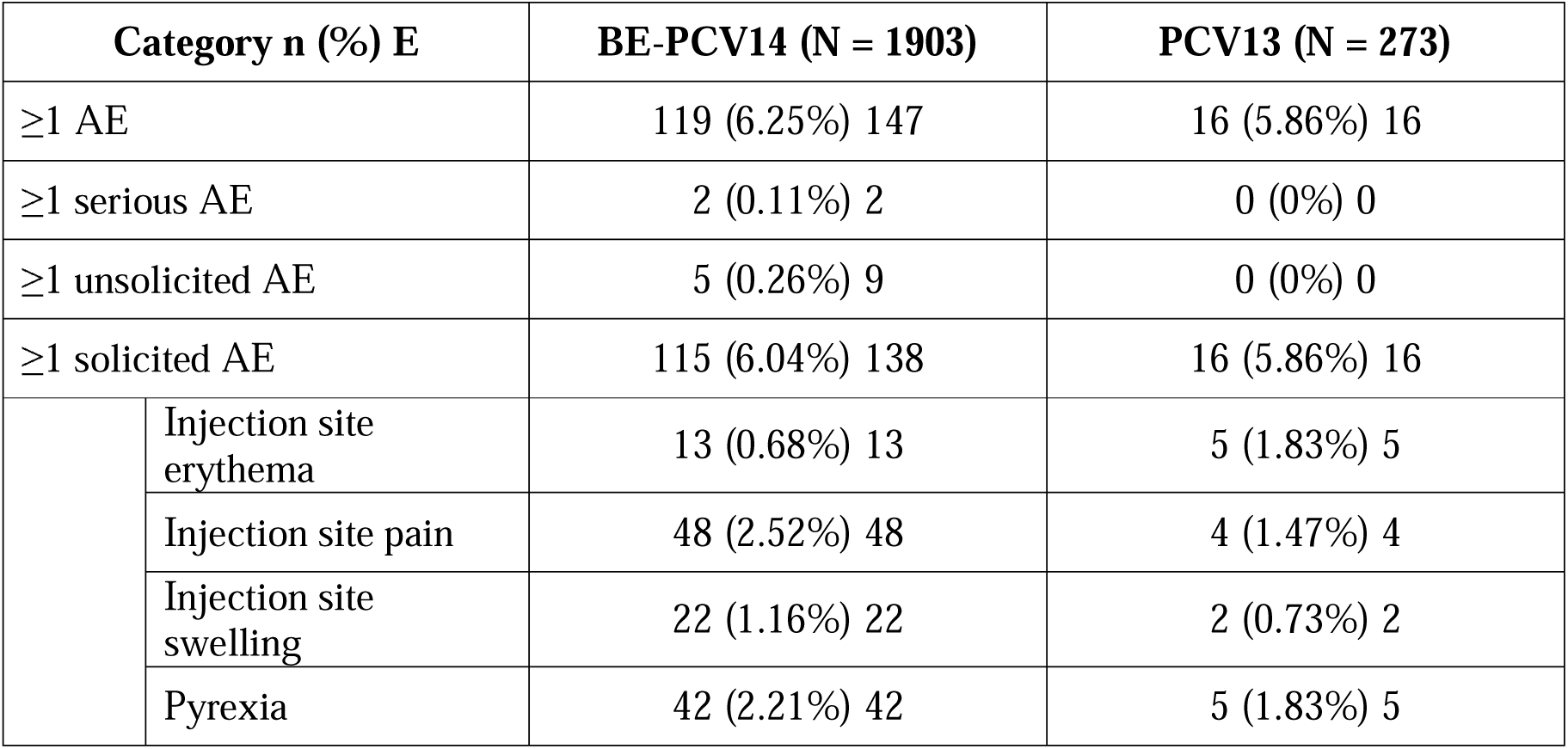
Adverse events (AEs) after a booster dose of BE-PCV14 or PCV13.

| Category n (%) E |  | BE-PCV14 (N = 1903) | PCV13 (N = 273) |
| --- | --- | --- | --- |
| ≥1 AE |  | 119 (6.25%) 147 | 16 (5.86%) 16 |
| ≥1 serious AE |  | 2 (0.11%) 2 | 0 (0%) 0 |
| ≥1 unsolicited AE |  | 5 (0.26%) 9 | 0 (0%) 0 |
| ≥1 solicited AE |  | 115 (6.04%) 138 | 16 (5.86%) 16 |
|  | Injection site erythema | 13 (0.68%) 13 | 5 (1.83%) 5 |
|  | Injection site pain | 48 (2.52%) 48 | 4 (1.47%) 4 |
|  | Injection site swelling | 22 (1.16%) 22 | 2 (0.73%) 2 |
|  | Pyrexia | 42 (2.21%) 42 | 5 (1.83%) 5 |

### Immunogenicity findings

#### BE-PCV14 induced a robust immunogenic response when administered as a booster dose after a three-dose primary series

The booster objectives of this Phase IV study included assessing the immunogenicity of a BE-PCV14 booster dose administered to toddlers aged 12–15 months of age after they had received a three-dose infant primary series. To answer this question, GMC ratios of post-booster to pre-booster dose and post-booster to post-primary doses for all 14 serotypes and cross-reactive serotype 6A were calculated (Table 3).

**Table 3:** BE-PCV14 Post-booster / pre-booster and post-booster / post-primary GMC ratios.

| Serotype | GMC Ratio (95%CI) |  |
| --- | --- | --- |
|  | Post-booster / Pre-booster | Post-booster / Post-primary |
| 1 | 4.42<br>(2.17, 8.98) | 1.97<br>(1.14, 3.40) |
| 3 | 2.77<br>(1.43, 5.34) | 1.51<br>(0.91, 2.49) |
| 4 | 3.59<br>(1.76, 7.30) | 1.47<br>(0.86, 2.50) |
| 5 | 2.91<br>(1.35, 6.30) | 0.96<br>(0.54, 1.72) |
| 6A | 3.11<br>(1.32, 7.35) | 4.09<br>(2.09, 8.00) |
| 6B | 3.57<br>(1.60, 7.96) | 3.60<br>(1.84, 7.03) |
| 7F | 3.23<br>(1.58, 6.61) | 1.18<br>(0.67, 2.06) |
| 9V | 4.13<br>(1.87, 9.13) | 1.49<br>(0.81, 2.73) |
| 14 | 2.38<br>(1.12, 5.05) | 1.68<br>(0.98, 2.91) |
| 18C | 3.94<br>(1.69, 9.21) | 1.60<br>(0.88, 2.93) |
| 19A | 4.70<br>(2.15, 10.27) | 2.26<br>(1.27, 4.00) |
| 19F | 5.31<br>(2.38, 11.84) | 2.30<br>(1.25, 4.23) |
| 22F | 4.06<br>(1.95, 8.43) | 2.11<br>(1.15, 3.88) |
| 23F | 3.63<br>(1.58, 8.38) | 2.54<br>(1.37, 4.71) |
| 33F | 3.15<br>(1.39, 7.14) | 2.99<br>(1.54, 5.80) |

GMC = Geometric Mean Concentration, CI = Confidence Interval

Post-booster / pre-booster dose GMC ratios ranged from ∼2.4 for serotype 14 to >5 for serotype 19F. Importantly, the two additional serotypes included in BE-PCV14 demonstrated strong booster responses with a >4-fold rise in GMC for serotype 22F and >3-fold rise in GMC for serotype 33F. For Post-booster / post-primary dose GMC ratios ranged from ∼1 for serotype 5 to >4 for serotype 6A.

To further understand the time-course of serotype-specific IgG antibody levels, the post-primary, pre-booster, and post-booster GMCs of children who received either BE-PCV14 or PCV13 were plotted (Figure 2). The GMCs at different timepoints were comparable in children who received BE-PCV14 or PCV13. A strong booster response for serotypes 22F and serotype 33F, which are present only in BE-PCV14, and for cross-reactive serotype 6A was noted.

**Figure 2:**
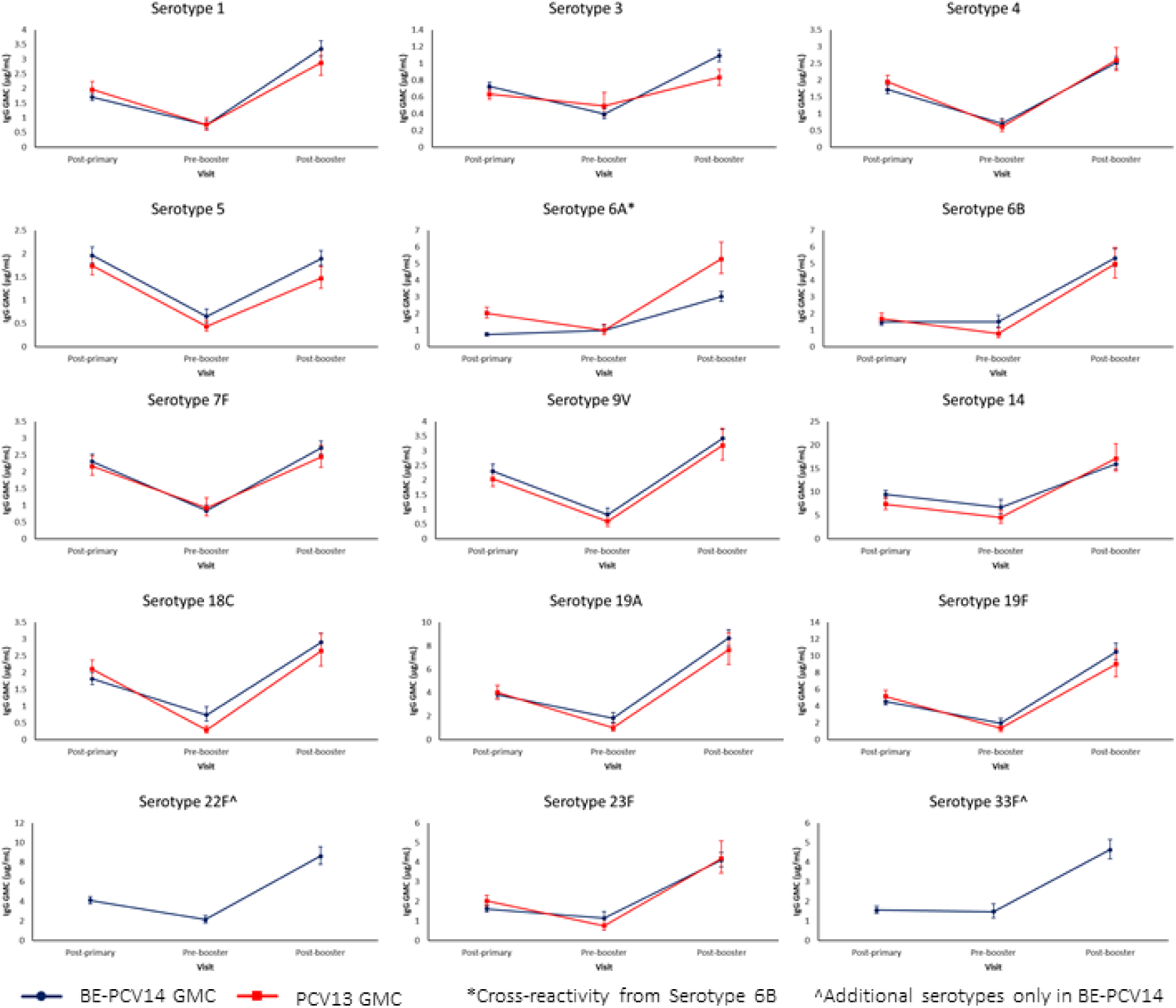
Post-primary, pre-booster, and post-booster GMCs after BE-PCV14 or PCV13.

**Figure 3:**
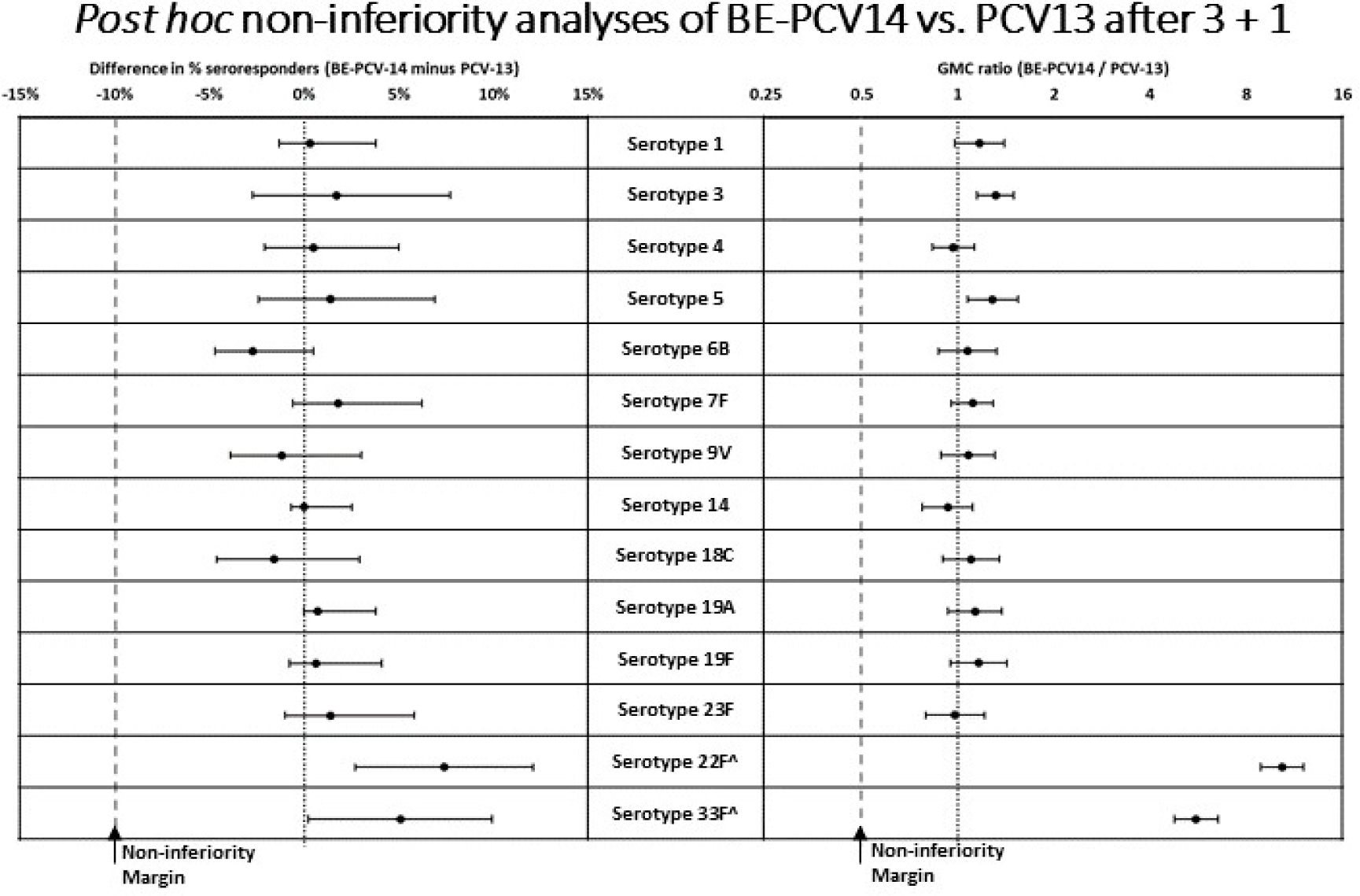
*Post hoc* non-inferiority analyses of BE-PCV14 to PCV13 after 3p+1. ^Additional serotypes 22F and 33F were compared to the worst performing serotype 3 of PCV13

#### Post-booster seroresponse rates and GMCs after BE-PCV14 or PCV13 were comparable

The booster responses were also assessed by seroresponse rates (SRRs) defined as the percentage of study participants with serotype-specific IgG antibody levels ≥0.35 µg/mL. BE-PCV14 induced high SRRs for all 14 serotypes and cross-reactive serotype 6A. The SRRs for all 15 serotypes tested were above 90% ranging from ∼93% for serotype 3 to 100% for serotypes 14 and 19A. These were comparable to the SRRs after PCV13 for the shared serotypes (ranging from ∼91% for serotype 3 to 100% for serotype 14). Similarly, post-booster GMCs after BE-PCV14 and PCV13 were comparable for shared serotypes (Table 4).

**Table 4:**
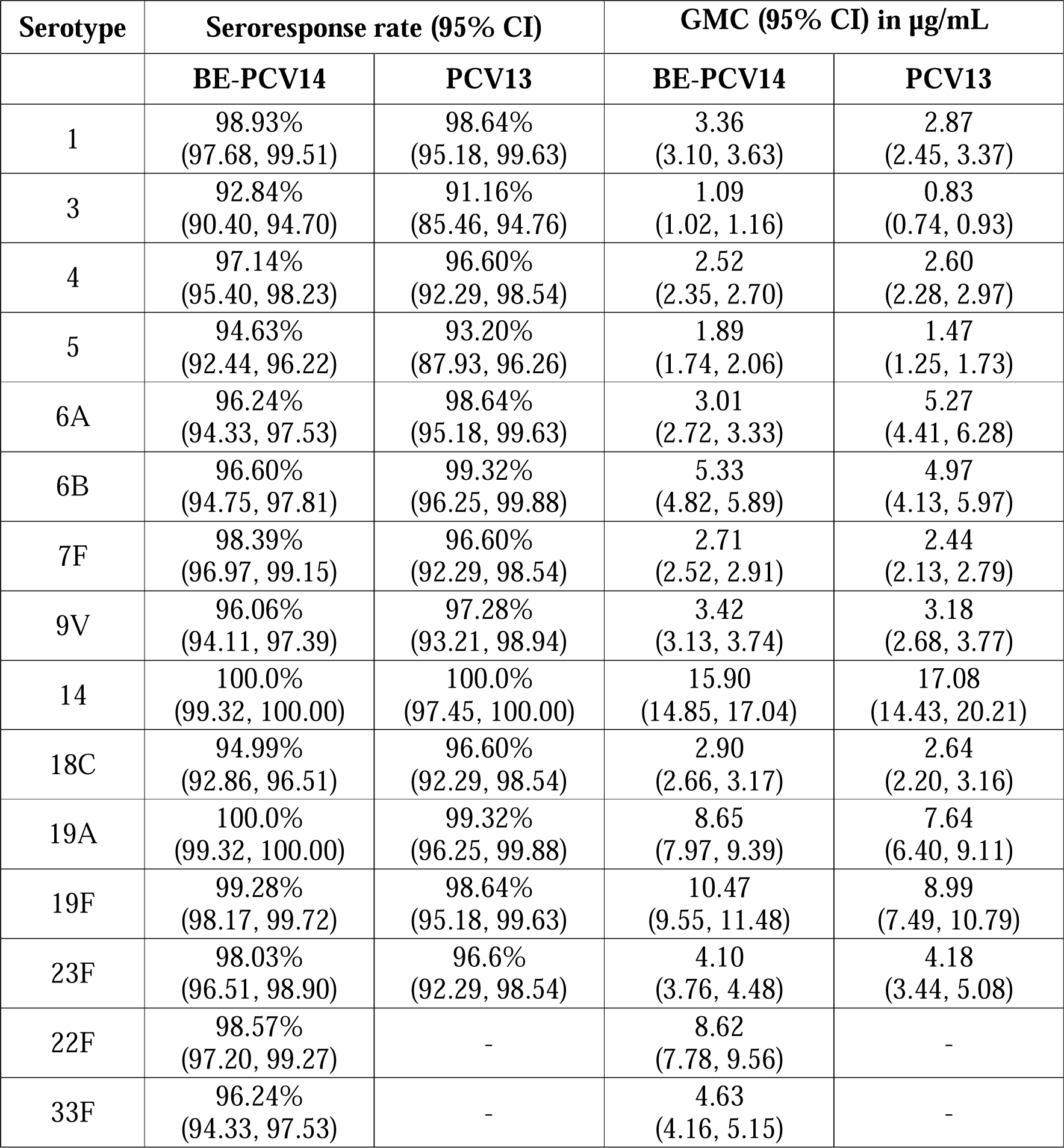
Post-booster seroresponse rates and GMCs after BE-PCV14 or PCV13.

#### Seroresponse rates and GMCs post-booster after BE-PCV14 were non-inferior to PCV13 in post hoc analyses

To further compare the post-booster responses of BE-PCV14 with PCV13, *post hoc* non-inferiority analyses were performed on both the difference in the percentage of seroresponders and the GMC ratio for each of the 14 serotypes. Non-inferiority was to be concluded if the lower limit of the 95% confidence interval (CI) for the difference in the percentage of seroresponders of BE-PCV14 and PCV13 was > -10%. Similarly, non-inferiority was to be concluded if the lower limit of the 95% confidence interval of the post-booster GMC ratio of BE-PCV14 to PCV13 was >0.5. By both these metrics, BE-PCV14 was non-inferior to PCV13 for all 14 serotypes. Importantly, the 95% CIs for all 12 shared serotypes either included or were greater than the reference for the difference in seroresponse and GMC ratios (reference for difference – 0%, reference for GMC ratio – 1).

## Discussion

The immunogenicity and safety profile of BE-PCV14 have been previously demonstrated **(11,12,13)** in 2p+1 and 3p+0 immunization schedules. The current prospective, multicentre, phase IV study evaluated the safety and immunogenicity of four 0.5 mL intramuscular doses (in a 3p+1 schedule) of BE-PCV14 in healthy Indian infants and toddlers. The overall safety profile of a booster of a BE-PCV14 was comparable to that of PCV13. The majority of local or systemic AEs reported up to 28 days after the booster dose in the current study were predominantly mild in severity. Injection-site and pyrexia were the most commonly reported local and systemic solicited AEs, respectively. The safety results presented in this study extend the findings of our previous phase III studies. These results are in line with other studies of BE-PCV14 **(11,12)**, and those conducted for PCV13 and PCV15 **(17, 18)**. Overall, a BE-PCV14 booster dose was generally well tolerated in toddlers.

The current study also evaluated the immunogenicity of the 3p+1 schedule of BE-PCV14 in comparison to PCV13. BE-PCV14 induced a robust immunogenic response when administered as a booster dose after a three-dose primary series. The post-booster / pre-booster dose GMC ratios ranged from ∼2.4 to >5. In addition, the two additional serotypes included in BE-PCV14 demonstrated strong booster responses with a >4-fold rise in GMC for serotype 22F and >3-fold rise in GMC for serotype 33F. The post-booster / post-primary dose GMC ratios ranged from ∼1 to >4. The SRRs for all 15 serotypes tested were above 90% (∼93% to 100%) and were comparable to PCV13 for the shared serotypes (∼91% to 100%). In totality, a booster dose of BE-PCV14 was immunogenic for all 14 serotypes included in the vaccine, including the two new emerging serotypes 22F and 33F. Additionally, as has now been shown in three other Phase III trials **(11,12,13)**, the inclusion of serotype 6B in BE-PCV14 induces cross-protective antibodies against serotype 6A further expanding the spectrum of protection offered by BE-PCV14. Compared with studies of PCV13 and PCV15, BE-PCV14 induces stronger immune responses either by expanding coverage compared to PCV13 or by inducing higher GMCs than PCV15 **(18)**. Taur et al. **(19)** compared PCV13 in a 3p+1 schedule against PCV14-BE in a 3p+0 schedule, a structurally biased setup because it embeds a booster advantage for PCV13 by design. Data described in this manuscript show the BE-PCV14 is immunologically non-inferior to PCV13 after completion of a 3p+1 series, consistent with the already published non-inferiority of BE-PCV14 vs PCV13 in the pivotal infant trial schedule **(11)**. Accordingly, the model’s large downward “effectiveness adjustment” applied to BE-PCV14 is not justified when the vaccines are evaluated on comparable, booster-containing regimens, undermining the claimed dominance of PCV13 in their model. In fact, with a lower private market price, and an expanded serotype coverage compared with PCV13, BE-PCV14 would render PCV13 a dominated strategy (i.e., higher costs and fewer health gains) if their own model were to be rerun!

Another Indian study compared the immunogenicity and safety of PCV13 and PCV7 in healthy infants receiving either of the study vaccines in a 3p+1 schedule at 6, 10, and 14 weeks of age followed by a booster dose at 12 months of age, along with routine pediatric vaccinations. IgG responses against the vaccine serotypes evaluated 1 month after the last dose of the primary series and after the booster dose showed comparable results between the vaccine groups, along with a tolerable safety profile **(20)**.

A recent WHO position paper on PCVs in children presented an analysis from the PSERENADE study that showed no evidence of significant differences in the decline of vaccine-type IPD by infant schedule **(21)**. However, multiple efficacy and effectiveness studies have demonstrated the benefit of the 3p+1 schedule for PCVs. Whitney et al. found very high real-world effectiveness of PCV7 against vaccine-serotype invasive pneumococcal disease (96% in healthy children and 81% in those with comorbidities) and protection even against penicillin-susceptible disease, supporting strong impact under routine multi-dose use **(22)**. A review describes how PCV13 introduction (including settings using a traditional 3p+1 schedule) was followed by marked declines in pediatric pneumococcal outcomes (IPD, pneumonia/AOM, and carriage) with noted attention to emerging non-vaccine serotypes **(23)**. In a Finnish randomized trial, PCV7 led to significant reductions in pneumococcal AOM (34%) and vaccine-serotype AOM (57%), including similar effectiveness for serotype 6A, highlighting the value of priming against vaccine serotypes **(24)**. Lalwani et al. directly provide data supporting the use of PCV boosters in Indian children—evidence relevant when justifying a 3p+1 schedule for BE-PCV14 **(25)**. Importantly, Whitney et al. went on to demonstrate that the inclusion of serotype 6B in PCV7 produces a cross-protection of ∼76% against serotype 6A. BE-PCV14 has a higher concentration of serotype 6B than PCV7 per vaccine dose, and produces significantly higher GMCs as well.

### Limitations

Our study had a few limitations. Non-inferiority passing criteria were not set *a priori*. However, *post hoc* analyses utilizing standardized criteria across PCV trials demonstrated non-inferiority of BE-PCV14 to PCV13 for all 14 serotypes by both post-booster seroconversion rates and IgG geometric mean concentrations. Additionally, the pivotal Phase III trial of BE-PCV14 completed in 2022 was specifically powered to demonstrate immunogenic non-inferiority to PCV13. The booster immunogenicity analyses were initially planned for infants who had been randomized to receive either BE-PCV14 (N=300) or PCV13 (N=300). Following regulatory guidance, the analyses for BE-PCV14 were expanded to include additional infants from a non-randomized arm as well (total of N=600). In addition to these limitations, the absence of opsonophagocytic assay (OPA) was also a limitation of this study. Immunogenic evaluation was based on Pneumococcal capsular polysaccharide (PnCPS) IgG concentrations and it did not include functional antibody activity via OPA, which is a better correlate of protection for PCVs. However, BE-PCV14’s phase III pivotal study demonstrated comparable OPA titters with PCV13 **(11)**.

### Conclusions

As part of a large post-marketing study of PNEUBEVAX 14^®^ (BE-PCV14) that led to WHO prequalification **(26)**, we demonstrate robust immunogenicity in a 3p+1 schedule (primary series at 6, 10, 14 weeks and booster at 12 –15 months of age) with an acceptable safety profile in Indian infants. The immunogenicity, safety, and tolerability profiles of BE-PCV14 were comparable to PCV13 when co-administered with routine pediatric immunizations. It also induces strong cross-protective immune responses against serotype 6A.

## Data Availability

All data produced in the present work are contained in the manuscript.

## Notes

### Competing Interest Statement

ST, RVM, SG, SN, VY, RRM, CD, KT, SD, PP, RE, and SND are or were employees of Biological E. Limited and do not own any equity in the company. All the other authors were principal investigators that received funding from Biological E. Limited to conduct the study.

### Clinical Trial

CTRI/2023/09/057894

### Funding Statement

This study was funded by Biological E. Limited.

### Author Declarations

Ethical approval was obtained from the following IECs/IRBs: IEC Aditya Multispeciality Hospital; Institutional Ethics Committee- All India Institute of Medical Sciences, BBSR; Institutional Human Ethics Committee All India Institute Of Medical Sciences; Institute Ethics Committee- AIIMS- Raipur; Institutional Ethics Committee for Clinical Trial All India Institute of Medical sciences; Ethics Committee of Bangalore Medical College and Research Institute; BAPS Hospital Institutional Ethics Committee; Istitutional Ethics Committee- Belagavi Institute Of Medical Sciences; Institutional Ethics Committe, BGS Global Institute of Medical Sciencese; Institutional Human Ethics Committee- Chettinad Hospital And Research Institute; IEC Chirayu Hospital; Institutional EthicsCommittee D Y Patil Medical College; Institutional Ethics Committee for ESIC; Gillurkar Hospital Ethics Committee; Institutional Ethics Committee Government Medical College; Guru Teg Bahadur Hospital Ethics Committee; Jehangir Clinical Development Cente Pvt.Ltd; Institutional Ethics Committee, Jawahar Lal Nehru Medical College; Institutional Ethics Committee, JSS Medical College, JSS Hospital; KEM Hospital Research Centre Ethics Committee; IEC King George Hospital; Institutional Ethics Committee, KLE University; Institutional Ethics Committee NKP Salve Institute of Medical Sciences; IEC, Maharaja Agrasen Hospital; Institutional Ethics Committee-MGIMS; Penta-Med Ethics Committee-Medipoint Hospitals Pvt. Ltd; Medstar Speciality hospital Ethics Committee; Institutional Ethics Committee-Leelamani Hospital; PMCHRI-IHEC; Institutional Ethics Committee, PGIMS; Institutional Ethics Committee, Rajarajeshwari Medical College and Hospital; Shubham Sudbhawana Superspeciality Hospital Ethics Committee; Institutional Ethics Committee for Human Research, Medical College; Ethics committee St Theresas Hospital.

