## Supplementary Information for "Safety and Immunogenicity of Biological E’s 14-valent Pneumococcal Conjugate Vaccine (PNEUBEVAX 14^®^) Administered in a 3p+1 Schedule to Healthy Indian Infants and Toddlers: A Prospective, Multicenter, Active Controlled Phase IV Trial"

**Appendix 1: Eligibility criteria**

**Inclusion Criteria:**

Participants who had met all the following inclusion criteria were selected for study participation:

1. Healthy pneumococcal conjugate vaccine-naïve (PCV-naive) infants as established by medical history and clinical assessment before entering into the study. PCV-naïve infants are those who have not been previously vaccinated with any licensed or investigational pneumococcal vaccine (only for Safety arm).
2. Healthy pneumococcal conjugate vaccine-naïve (PCV-naive), Pentavalent vaccine (DTwP-rHepB-Hib) naïve, IPV vaccine naïve and live attenuated Rotavirus vaccine naïve infants as established by medical history and clinical assessment before entering into the study. PCV/Pentavalent/IPV/Rotavirus vaccine-naïve infants are those who have not been previously vaccinated with any licensed or investigational PCV/Pentavalent/IPV/Rotavirus vaccines (only for immunogenicity arm).
3. Infants between 6-8 weeks of age (42-56 days, both days inclusive) of either gender, at the time of first dose of vaccination.
4. Healthy Infants with body weight ≥ 3300 gms at the time of screening.
5. Participants’ parent(s)/ LAR(s) who, in the opinion of the investigator, can and will comply, with the requirements of the protocol (e.g. completion of the diary cards, return for follow-up visits, with access to a consistent means of telephone contact, either residential landline or mobile).
6. Participant’s parent(s)/LAR(s) willing to provide written or thumb printed informed consent (including audio visual recording of consent process) prior to performing any study specific procedure
7. Infants with a minimal vaccination status for their age at the time of enrolment (“minimal” defined as single dose of only BCG, Hepatitis B &/or Polio vaccine prior to enrolment).

**Exclusion Criteria**

Participants who had met any of the following exclusion criteria were not enrolled in the study:

1. Child in care, defined as a child who has been placed under the control or protection of an agency, organisation, institution or entity by the courts, the government or a government body, acting in accordance with powers conferred on them by law or regulation. The definition of a child in care can include a child cared for by foster parents or living in a care home or institution, provided that the arrangement falls within the definition above. The definition of a child in care does not include a child who was adopted or has an appointed legal guardian.
2. Evidence of previous Streptococcus pneumoniae infection or pneumococcal vaccination.
3. Evidence of previous or intercurrent or known exposure to diphtheria, tetanus, pertussis, hepatitis B, poliomyelitis, H. influenzae type b and rotavirus diseases (immunogenicity arm only)
4. Use of any investigational or non-registered product (drug or vaccine) during the period starting 30 days before the administration of study vaccine (Day -29 to Day 0), or planned use during the study period other than the study vaccine.
5. Any medical condition that in the judgment of the investigator would make intramuscular injection unsafe (eg. coagulation abnormalities).
6. Concurrently participating in another clinical study, at any time during the study period, in which the participant has been or was exposed to an investigational or a non-investigational vaccine/product (pharmaceutical product or device).
7. History of allergic disease or history of a serious reaction to any prior vaccination or known hypersensitivity likely to be exacerbated by any component of the study vaccines.
8. History of any neurological disorders, meningitis or seizures.
9. Infant who has had a sibling die of sudden infant death syndrome (SIDS) or die suddenly and without apparent other cause or preceding illness in the first year of life.
10. Infant is a direct descendant (child or grand-child) of any person employed by the Sponsor, the Contract Research Organization (CRO) or the Study Site (including the PI and study site personnel).
11. Acute disease and/or fever at the time of vaccination.

- Fever was defined as the endogenous elevation of at least one measured body temperature of ≥ 38◦C (≥ 100.4◦F).

12. Acute or chronic, clinically significant pulmonary, cardiovascular, hepatic or renal functional abnormality, as determined by physical examination and Principal investigator judgement.

**Appendix 2: SAATHI-14 Consortium principal investigators and sites**

| **#** | **Principal Investigator** | **Site Name, City, State** | **IEC Name** |
| --- | --- | --- | --- |
| 1 | Dr Bhagirathi Dwibedi | All India Institute of Medical Sciences, Khordha, Orissa | Institutional Ethics Committee- All India Institute of Medical Sciences, BBSR |
| 2 | Dr Hira Lal Bhalla | All India Institute Of Medical Sciences, Gorakhpur, Uttar Pradesh | Institutional Human Ethics Committee All India Institute Of Medical Sciences |
| 3 | Dr Atul Jindal | All India Institute of Medical Sciences (AIIMS), Raipur, Chhattisgarh | Institute Ethics Committee- AIIMS- Raipur |
| 4 | Dr Urmila Chauhan | All India Institute of Medical Sciences (AIIMS), Nagpur, Maharashtra | Institutional Ethics Committee for Clinical Trial All India Institute of Medical sciences |
| 5 | Dr Malesh K | Bangalore Medical College and Research Institute, Bangalore, Karnataka | Ethics Committee of Bangalore Medical College and Research Institute |
| 6 | Dr Kiritkumar Jesabhai Sisodia | BAPS Pramukh Swami Hospital, Surat, Gujarat | BAPS Hospital Institutional Ethics Committee |
| 7 | Dr Chinmayi Joshi | Belgavi Institute of Medical Sciences, Belgaum, Karnataka | Istitutional Ethics Committee- Belagavi Institute Of Medical Sciences |
| 8 | Dr Ramesh M | BGS Global Institute of Medical Sciences, Bangalore, Karnataka | Institutional Ethics Committe, BGS Global Institute of Medical Sciencese |
| 9 | Dr M Alexander | Chettinad Hospital And Research Institute, Chennai, Tamil Nadu | Institutional Human Ethics Committee- Chettinad Hospital And Research Institute |
| 10 | Dr Kishori Sharan Agarwal | Chirayu Hospital, Jaipur, Rajasthan | IEC Chirayu Hospital |
| 11 | Dr Saxena Amit Shyammohan | D. Y. Patil Hospital and Research centre, Pune, Maharashtra | Institutional EthicsCommittee D Y Patil Medical College |
| 12 | Dr Anil Kumar Pandey | ESIC Medical College & Hospital, Faridabad, Haryana | Institutional Ethics Committee for ESIC |
| 13 | Dr Girish P Charde | Gillurkar Multispeciality Hospital, Nagpur, Maharashtra | Gillurkar Hospital Ethics Committee |
| 14 | Dr Prabha Khaire | Government Medical College & Hospital, Aurangabad, Maharashtra | Institutional Ethics Committee Government Medical College |
| 15 | Dr Manish Narang | GTB Hospital, North East, Delhi | Guru Teg Bahadur Hospital Ethics Committee |
| 16 | Dr Bafna Sanjay Mohanlal | Jehangir Clinical Development Centre Pvt. Ltd., Pune, Maharashtra | Jehangir Clinical Development Cente Pvt.Ltd |
| 17 | Dr Jai Prakash Narayan | JLN Medical College, Ajmer, Rajasthan | Institutional Ethics Committee, Jawahar Lal Nehru Medical College |
| 18 | Dr MD Ravi | JSS Hospital, Mysore, Karnataka | Institutional Ethics Committee, JSS Medical College, JSS Hospital |
| 19 | Dr Anand Shantaram Kawade | KEM Hospital Research Centre, Pune, Maharashtra | KEM Hospital Research Centre Ethics Committee |
| 20 | Dr BS Chakravarthy | King George Hospital, Visakhapatnam, Andhra Pradesh | IEC King George Hospital |
| 21 | Dr N S Mahantshetti | KLES Dr. Prabhakar Kore Hospital & Medical Research Centre, Belgaum, Karnataka | Institutional Ethics Committee, KLE University |
| 22 | Dr Anju JK Mehrotra | Lata Mangeshkar Hosptial, Nagpur, Maharashtra | Institutional Ethics Committee NKP Salve Institute of Medical Sciences |
| 23 | Dr Vinay Kumar Gill | Maharaha Aagrasen Superspeciality Hospital, Jaipur, Rajasthan | IEC, Maharaja Agrasen Hospital |
| 24 | Dr Bishan Swarup Garg | Mahatma Gandhi Institute of Medical Sciences (MGIMS), Wardha, Maharashtra | Institutional Ethics Committee-MGIMS |
| 25 | Dr Jog Pramod Prabhakar | Medipoint Hospital, Pune, Maharashtra | Penta-Med Ethics Committee-Medipoint Hospitals Pvt. Ltd |
| 26 | Dr Prashanth MV | Medstar Speciality Hospital, Bangalore, Karnataka | Medstar Speciality hospital Ethics Committee |
| 27 | Dr Tripathi Virendra Nath | New Leelamani Hospital, Kanpur Nagar, Uttar Pradesh | Institutional Ethics Committee-Leelamani Hospital |
| 28 | Dr Ramanath Andy Karayar | Panimalar Medical College Hospital & Research Institute, Chennai, Tamil Nadu | PMCHRI-IHEC |
| 29 | Dr B Sudhakar | Priya Childrens Hospital, Krishna, Andhra Pradesh | Institutional Ethics Committee Anu Hospitals, Vijayawada,  Andhra Pradesh |
| 30 | Dr Savita Verma | Pt. BD Sharma PGIMS, Rohtak, Haryana | Institutional Ethics Committee, PGIMS |
| 31 | Dr Prema | Rajarajeswari Medical College and Hospital, Bangalore, Karnataka | Institutional Ethics Committee, Rajarajeshwari Medical College and Hospital |
| 32 | Dr Madhukar Pandey | Shubham Sudbhawana Superspeciality Hospital, Varanasi, Uttar Pradesh | Shubham Sudbhawana Superspeciality Hospital Ethics Committee |
| 33 | Dr Pareshkumar A Thakkar | SSG General Hospital, Vadodara, Gujarat | Institutional Ethics Committee for Human Research, Medical College |
| 34 | Dr G Balakishor | St. Theresas Hospital, Hyderabad, Telangana | Ethics committee St Theresas Hospital |
| 35 | Dr Nagarjuna Chadalavada | Aditya multispeciality hospital, Guntur, Andhra Pradesh | IEC Aditya Multispeciality Hospital |
